# Neuroblastoma in Adults: A Scoping Review of Presentations, Genetics and Therapies

**DOI:** 10.1101/2024.05.22.24307615

**Authors:** Bader H. Alsaikhan, Basmah Alwahhabi, Abdullah Alshalan, Alex Koziarz, Abdullah M. Alkhayal, Khalid Alrabeeah

## Abstract

**Purpose:** As a scoping review, evaluate the literature on the presentations, genetics, and therapies for neuroblastoma in adult patients.

**Methods:** We searched four databases for studies reporting adults with neuroblastoma. Cohort studies, case series, and case reports were synthesized qualitatively. Progression-free and overall survival were compared amongst cohort studies.

**Results:** Of 2287 unique records, 136 studies published in 141 articles were included. A total of 679 patients were included. On review of individual patient-level data, the adrenal gland and retroperitoneum were the most common primary site (47.3%). *MYCN* was rarely amplified: seven studies reported zero patients with MYCN amplified, two studies with a single patient, and one study with 3/7 patients. Adult patients appear to show a high frequency of somatic mutations, specifically *ALK* (42%) and *ATRX* (58%). Registry data of included studies showed 5- year overall survival to be 36.3% in adults aged ≥20 years.

**Conclusion:** Of nearly 700 cases of adult neuroblastoma published in the literature, the most common primary site is the adrenals or retroperitoneum. Relative to pediatric cases, adult cases demonstrate a considerable rate of somatic mutations such as *ALK* and *ATRX*. Registry data showed 5-year survival of 36%. Future studies evaluating targeted therapies in larger samples are needed.

**Take home messages:** 679 cases of adult neuroblastoma have been published in the literature.

Compared to pediatrics, adult cases have more somatic mutations (eg: ALK, ATRX).

Registry data showed that adult neuroblastoma has a 5-year survival of 36%.

## Introduction

Neuroblastoma is the most common extracranial solid tumor of childhood [1]. Unfortunately, over half of the children have metastatic disease on presentation. Neuroblastoma arises from cells of the neural crest that form the sympathetic ganglia and adrenal medulla. Tumors may occur anywhere along the sympathetic chain within the neck, thorax, retroperitoneum, or pelvis, or in the adrenal gland. The most common primary site includes the adrenal gland [1].

The occurrence of neuroblastoma is common in children however very rare in adults. Neuroblastoma is highly malignant, usually characterized by invasive growth and propensity for organ metastasis. The prognosis of adult patients with neuroblastoma is generally poor, and management is similar compared to the pediatric population.

The literature describing neuroblastoma is scattered with few cohort studies and countless case reports. Previously literature reviews include only a small fraction of the published cases, which may produce inaccurate summaries of the presentation, management, and prognosis. Therefore, we performed a scoping review with the aim to summarize all published reports of neuroblastoma in adults to inform clinicians of the available evidence of this heterogeneous disease.

## Methods

We performed this scoping review in an attempt to summarize the available literature on the presentations, genetics, and therapies for neuroblastoma in adult patients. All patients were required to be ≥18 years old and have a pathologic diagnosis of neuroblastoma. Studies were excluded if the primary site was intracranial, cutaneous, or in the extremities. We also excluded cases of olfactory neuroblastoma (esthesioneuroblastoma) and if only ganglioneuroblastoma were reported. If studies included a combination of neuroblastoma and ganglioneuroblastoma, we attempted to describe them separately. The following databases were searched for randomized controlled trials, observational studies, case series, and case reports: Medline, Embase, CINAHL, Web of Science. We used a combination of key word and medical subject headings such as ‘neuroblastoma’, ‘adult’, ‘geriatric’ from inception to 4 October 2022 (Medline strategy found in **Table A1**). There was no limitation on language or publication type (full text article or published conference abstract). The search was current until 7 October 2022.

References of all included studies were reviewed to identify studies not captured in the search of databases. Research ethics board approval was not required as all data was published and publicly available.

Studies were screened by a single investigator and reviewed by a second investigator for inclusion. All studies were reported qualitatively and categorized according to study design (cohort study, case series, case report). We aimed to identify the total number of published unique cases of adult neuroblastoma. We also aimed to report patient ages, genders, and primary tumor sites. For cohort studies with sufficient data, we reported relevant genetic abnormalities (eg, *MYCN* amplification, *ALK* or *ATRX* mutations), management strategies, and progression-free and overall survival.

In addition, we described a case of neuroblastoma in a 24-year-old male patient managed at King Abdullah University Hospital (Riyadh, Saudi Arabia).

## Results

### Scoping Review

#### Characteristics of included studies (number, study design, age, sex)

Our initial literature search identified 2287 unique articles, of which 101 were included following full text review. Forty articles were identified through manual searching of references of included articles. Accordingly, a total of 136 studies published in 141 articles were included in the scoping review (**Figure 1**; **Table 1**). **Table 2** details the genetic characteristics of neurroblastoma tumors of included studies. There were 6 cohort studies [2–7], 28 case series (**Table 3**) [11–38], and 102 case reports (**appendix Table A2**) [appendix references A1-A102].

**Figure 1:**
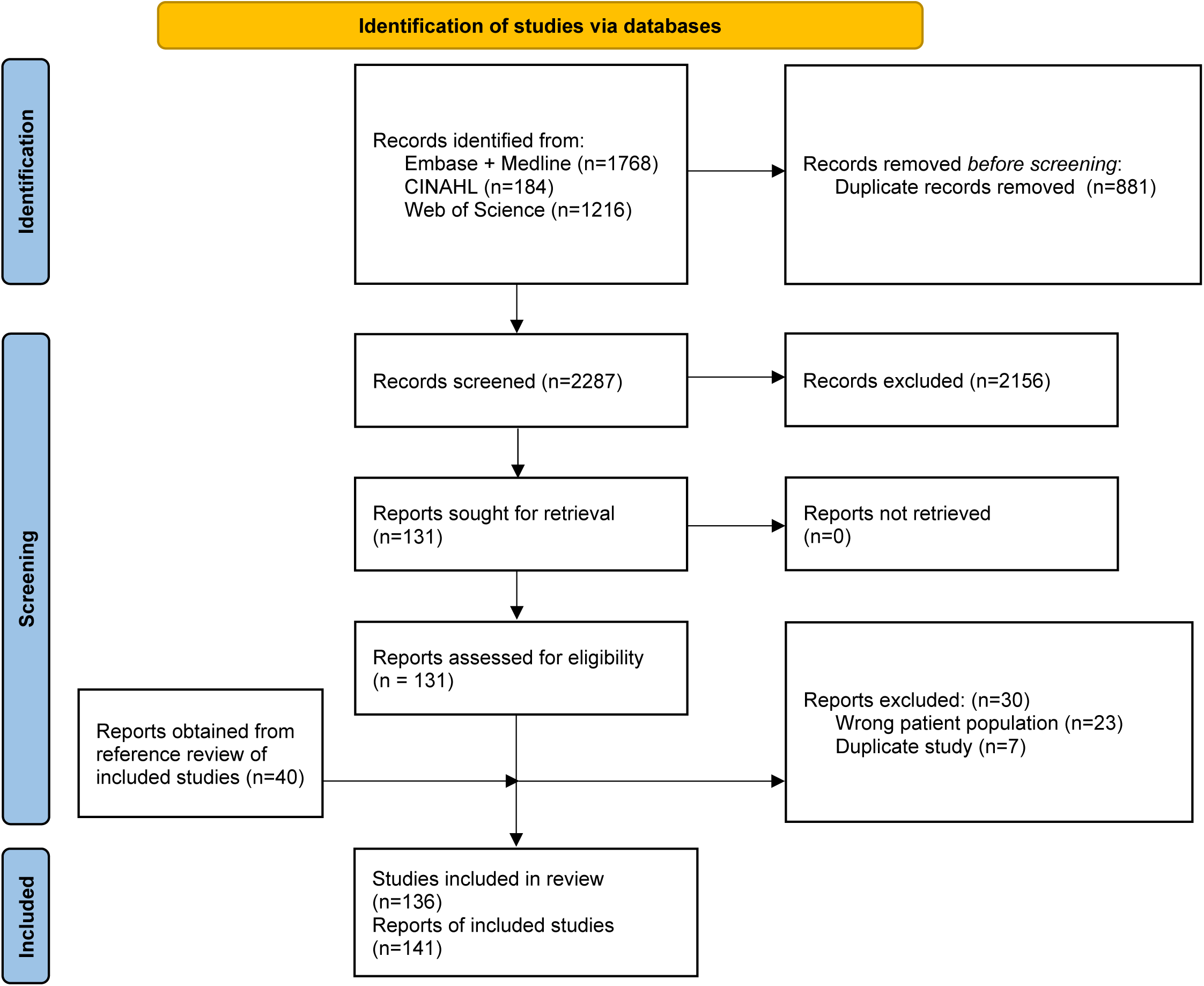
PRISMA flow diagram.

**Table 1:** Characteristics of 6 included cohort studies.

| Study | Country and Data Source | Patient cohort | Primary Tumor Site | Management | Outcome |
| --- | --- | --- | --- | --- | --- |
| Tang 1975 [2] | United States: Memorial Hospital 1948-1972 | N=27.<br>16 males (59%). | Mediastinum (7), retroperitoneum (5), pelvis (1), extremity (1), neck (1), multiple sites (12). | Surgical resection in 26 patients. Combined chemotherapy and radiation in 18. Radiation only in 7. | 67% 1-year survival. 28% 2-year survival. 12% lived over 5 years. |
| Polishchuk 2011 [3] | United States: University California San Francisco (recurrent or refractory cases) | N=16.<br>8 males (50%) | Abdomen (10), thorax (3), thoracoabdominal (1), pelvic (1), unknown (1). | <sup>131</sup>I-MIBG monotherapy | Median survival 36 months, estimated 3-year survival 43.2%. |
| Conter 2013 [4] | United States: MD Anderson Cancer Center 1994-2012 | N=118.<br>Mean age 47 (range 18-82) years. | NA | Specific management not available. | Adults similar survival to pediatrics (L1 p=0.40, L2 p=0.54, M p=0.73). L1 disease actuarial survival 94% at 3 years, 90% at 5 years, 69% at 10 years. L2 disease estimated survival 83% at 3 years, 73% at 5 years, 41% at 10 years. M actuarial survival 68% at 1 year, 33% at 2 years, 13% at 5 years. INRG stage was prognostic (p<0.001). |
| Rogowitz 2014 [5] | United States National Cancer Institute Surveillance, Epidemiology, and End-Results registry (SEER) | N=216.<br>35 patients >60 years (19%).<br>107 male (50%) | Most common site CNS (71) in patients 18-60 years. Most common site soft tissue including heart (21) in patients >60 years. | Specific management not available. | In patients >60 years, 5 year disease-specific survival was 40%. In patients with mediastinal, tracheal, and other resp neuroblastomas, 5 year disease-specific survival was 44% for 9 adult patients. |
| Sorrentino 2014 [6] | Italy: Istituto Giannina Gaslini | N=16.<br>Mean age 26 (range 18-37). 7 male (44%). | Abdomen (5), adrenal (5), thorax (2), pelvis (4).<br>6 patients metastatic disease on diagnosis. | **Stage 1/2 (4):**<br>All surgery, of which 1 also chemotherapy.<br>**Stage 3 (6):**<br>Various combinations of surgery (3), chemotherapy (4), radiation (2).<br>**Stage 4 (6):**<br>All chemotherapy, 3 also surgery and 2 radiation. | **Stage 1/2:**<br>2 died at 25 and 48 months. 1 alive with disease at 107 months. 1 remission at 130 months.<br>**Stage 3:**<br>5 patients died of disease at mean 70 months; 1 patient remission at 142 months.<br>**Stage 4:**<br>6 patients died of disease at mean 46 months. |
| Suzuki<br>2018 [7] | United States:<br>Memorial Sloan<br>Kettering Cancer<br>Center 1979-2015 | N=44.<br>Median age 25<br>(range 18-71).<br>25 males (57%). | Adrenal gland (26),<br>abdominal paraspinal<br>region (11), mediastinum<br>(5); bone or bone<br>marrow (2). | **Locoregional neuroblastoma (11):**<br>All surgery, and 8 additionally received chemotherapy.<br>**Stage 4 (33):**<br>27 high-dose chemotherapy. 1 patient crizotinib for ALK-mutation. Surgery was attempted in 11 patients.<br>Of 7 stage 4 patients with complete response, 5 received autologous stem cell transplant, 1 received autologous stem cell transplant and anti-GD2 immunotherapy with 3F8. 7 with incomplete response received anti-GD2 immunotherapy: 4 with m3F8 + GM-CSF, 2 with 3F8 alone, 1 with 3F8. | **Locoregional neuroblastoma:**<br>10-year progression-free 35.4% and overall survival was 61.4%<br>**Stage 4:**<br>7 (21%) achieved complete response (CR) after induction chemotherapy and/or surgery. 5-year progression-free survival 9.7%. 10- year overall survival 19%. Patients who achieved complete response after induction had superior survival. |

\*Three studies not detailed given overlapping patients [8-10]

**Table 2:** Genetic characteristics of neuroblastoma tumors in included studies.

| Study | Genetic testing | Genetic outcomes |
| --- | --- | --- |
| Polishchuk 2011 [3] | NA. | 0/9 MYCN amplifications. |
| Sorrentino 2014 [6] | NA. | 1/10 tested patients MYCN amplification.<br>1/5 tested patients 1p chromosome deletion. |
| Suzuki 2018 [7] | FISH; whole exome sequencing; DNA sequencing for *ALK*; targeted exome sequencing using Foundation One or MSK-IMPACT | 0/40 tumor *MYCN* amplification.<br>15/26 (58%) *ATRX* mutations or deletions.<br>10/16 (42%) *ALK* mutations.<br>Various somatic mutations in 11 patients with targeted exome sequencing including *BRCA1*, *IKZF2*, *NF2*, *MUTYH*, *TP53*, *CSF1R*, *CRLF2*, *FAT1*, *NUP93*, *BLM*, *PTPRD*, *RAC1*, *EZH2*, *SMARCA4*, *ERBB3*, *NOTCH3*, *PAK1*, *RBM10*, *BCL6*, *KIT*, *RASA1*. |
| Kushner 2003 [29] | NA. | 0/22 tumor *MYCN* amplification. |
| Podda 2010 [31] | NA. | 0/5 tumor *MYCN* amplification. |
| Berbegall 2014 [33] | aSNP, MLPA, and FISH in tumors with adequate DNA quality; FISH used for MYCN | 1/7 tumor heterogenous *MYCN* amplification.<br>1/6 17q gain.<br>1/6 11q deletion.<br>2/4 focal segmental chromosome aberration (+5p; +20p).<br>2/4 copy-neutral loss of heterozygosity (4q, 11p, 11p; 12q, 19p; 6p). |
| Mazzocco 2015 [36] | Double color FISH on interphase nuclei using *MYCN/LAF* probe for chromosomes 2p24 and 2q11; *ALK* assessed in exons 20 to 28 of the *ALK* gene; *ATRX* assessed in five exons (10, 16, 19, 30, 32) and five portions of exon 9 and flanking splice junctions of *ATRX* | 0/3 tumor *MYCN* amplification.<br>0/3 *ALK* mutations.<br>1/3 *ATRX* mutations (c.6572A>C p.D2191A).<br>1 patient 1p deletion, 9p deletion.<br>1 patient 2p gain.<br>1 patient 1p deletion, 9p deletion, 11q deletion, 17q gain. |
| Sartelet 2013 [32] | FISH for *MYCN* amplification. | 0/4 *MYCN* amplification. |
| Mahkamova 2014 [35] | NA. | 0/1 *MYCN* amplification. |
| Duan 2019 [37] | FISH used for MYCN copy number, chromosome 1p loss and 17q gain; | MYCN: 4/7 no amplification, 1/7 focal amplification, 1/7 diffuse amplification, 1/7 gain. |
|  | detecting alternate lengthening of telomeres (*ALT*); and for detecting telomerase gene *TERT* rearrangement. | 2/6 *ALK* mutation using IHC, of which no break detected using FISH.<br>4/5 *ATRX* mutation using IHC, of which 1 positive in 3.8% of cells using FISH.<br>1/5 1p deletion.<br>4/4 17q gain.<br>1/5 *TERT* rearrangement (diffuse amplification of 200x) detected using FISH. |

NA: not available; M: male; F: female; mo: month. MSK-IMPACT: Memorial Sloan Kettering Integrated Mutation Profiling of Actionable Cancer Targets; FISH: fluorescence in situ hybridization; IHC: immunohistochemistry;

**Table 3:**
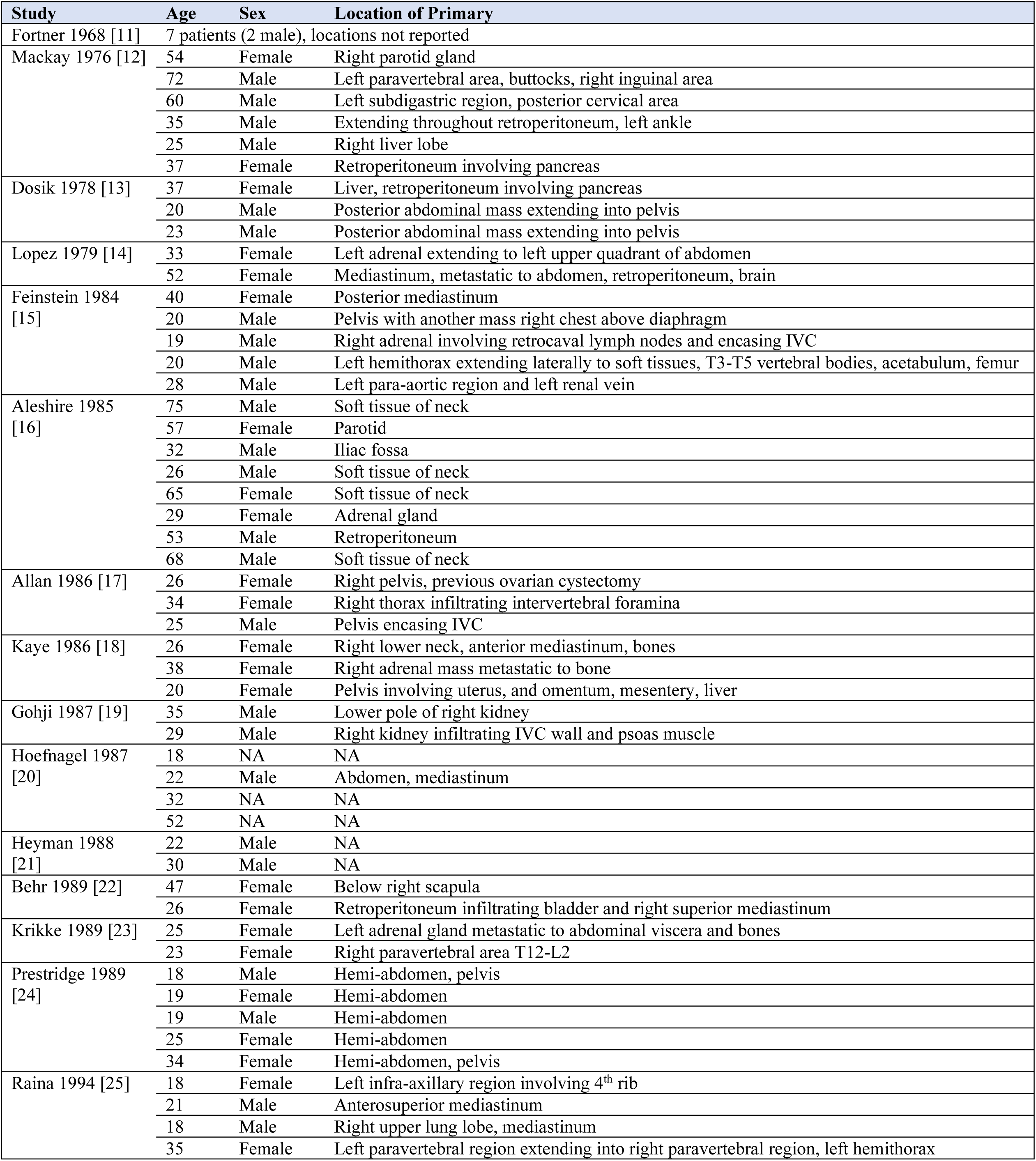
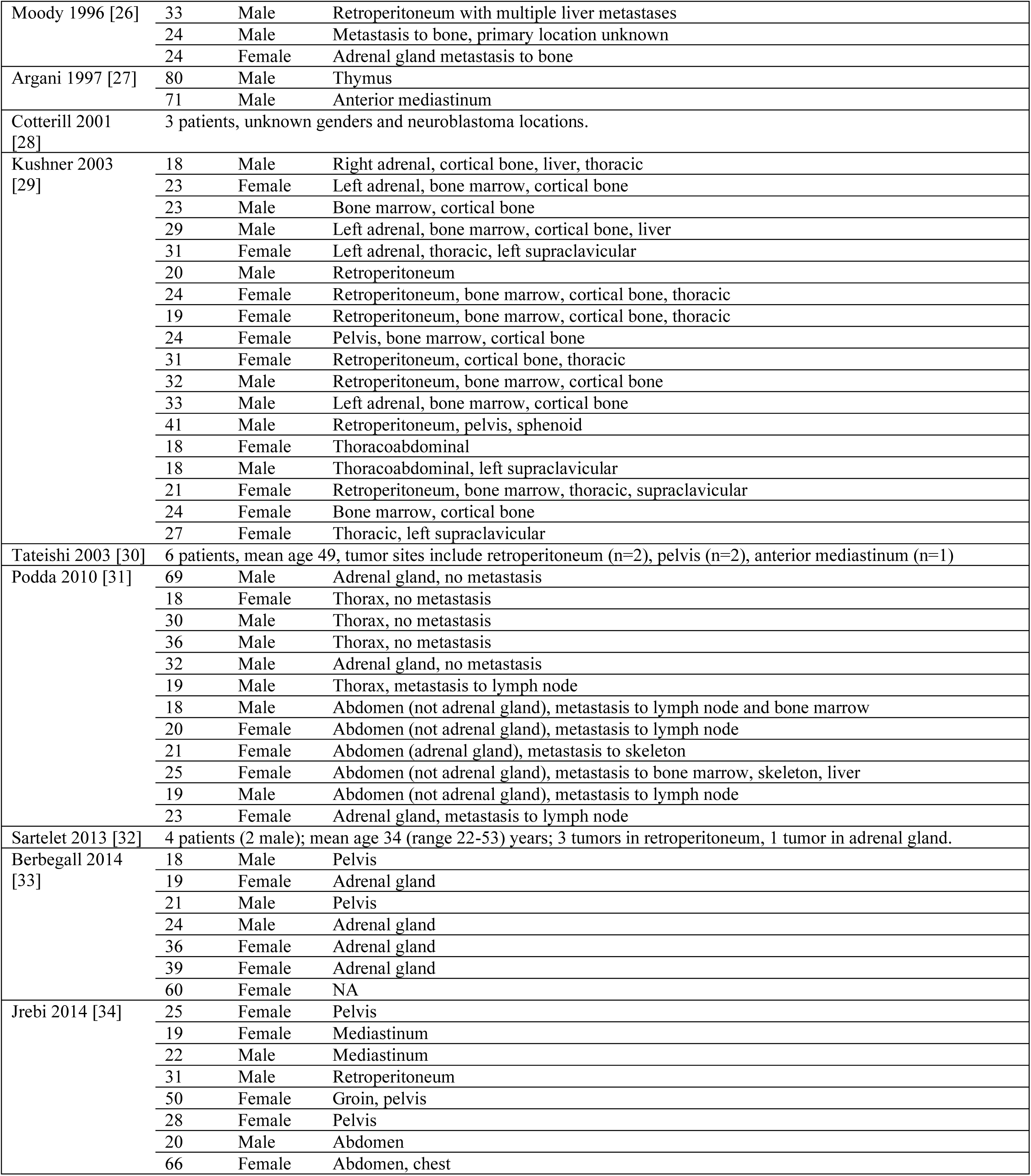
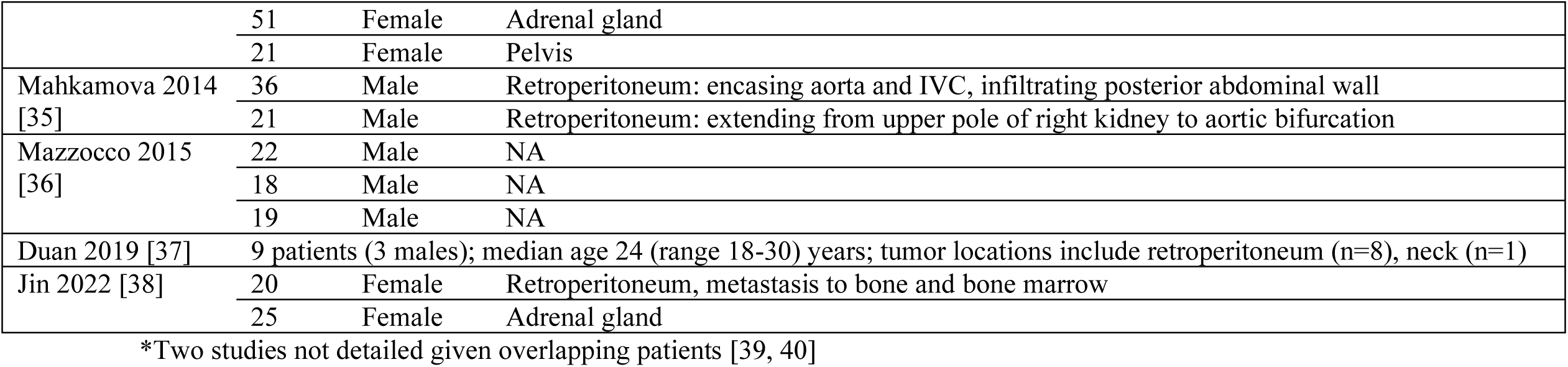
Characteristics of 28 included case series.

The total number of adult neuroblastoma cases was 679. Five articles included overlapping patients [8–10,39,40], but were still included as they provided different information. For cohort studies, male proportion ranged between 44% [6] to 59% [2]. Rogowitz and colleagues [5] reported the largest study, documenting 216 adult neuroblastoma cases sourced from the Surveillance, Epidemiology, and End-Results (SEER) registry (1973-2010). Of these 216 patients, 35 (16%) were over 60 years old.

#### Primary site of tumor

In the largest series [5], the most common primary tumor site in patients aged 18-60 years was the central nervous system (39%, not reported if this included sympathetic ganglia), followed by the retroperitoneum (17%), and endocrine tissues including thymus (14%). Suzuki and colleagues reported a series of 44 patients from Italy, whereby the most common primary site was adrenal gland (26/44, 59%), followed by the abdominal paraspinal region (11/44, 25%), and mediastinum (5/44, 11%). Tang and colleagues [2] reported a series of 27 patients published in 1975, of which 15 patients had a primary site identified. The most common primary site was the mediastinum (7/15, 47%), followed by the retroperitoneum (5/15, 33%). Notably, 12 patients were unable to have a primary site identified given multiple sites involved. Sorrentino and colleagues [6] report a series of 16 patients from Italy, of which the adrenal gland (5/16, 31%), and abdomen (5/16, 31%) were the commonest primary site, followed by the pelvis (4/16, 27%), and thorax (2/16, 13%). When totaling patients included in included case series (n=131), the most commonly reported site included the retroperitoneum not otherwise specified (n=38, 29%), followed by adrenal gland (n=24, 18%), pelvis (n=16, 12%), abdomen not otherwise specified (n=12, 9%), and mediastinum (n=11, 8%). If the primary sites of adrenal gland and retroperitoneum were combined, this site would total 47.3% (62/131) of patients.

#### Genetics

*MYCN* amplification occurred rarely in included studies of tested adult neuroblastoma patients: 0/22 patients in Kushner and colleagues [29], 0/5 in Podda and colleagues [31], 1/7 in Berbegall and colleagues [33], 0/40 in Suzuki and colleagues [7], 0/9 in Polishchuk and colleagues [3], 0/3 in Mazzocco and colleagues [36], and 1/10 in Sorrentino and colleagues [6]. The patient with *MYCN* amplification in Sorrentino and colleagues [6] was a 30-year-old female who presented with adrenal neuroblastoma with metastasis to the liver, and underwent radical tumor resection followed by cisplatin and three courses of peptichemio; she unfortunately died 10 months after initial presentation. Also in this study, one of five patients presented with 1p chromosome deletion. This was a 27-year-old male who presented with a neuroblastoma primary in the thorax with metastasis to bone. He underwent four courses of carboplatin and etoposide chemotherapy, and four courses of etoposide alone. He had a minor response but showed local and bone progression at 13 months and died 38 months following initial presentation.

Suzuki and colleagues reported a series of 44 patients [7], whereby somatic *ATRX* mutations or deletions were identified in 58% (15/26) of tested tumors and somatic *ALK* mutations were identified in 42% (10/16) of tested tumors. A proportion of this cohort underwent targeted exome sequencing at initial diagnosis (3/11) or relapse (8/11), which found a greater variety of mutations and total number of mutations at relapse. One patient had targeted exome sequencing performed both at initial diagnosis and at relapse: initial sequencing showed *CSF1R* V32G mutation, relapse sequencing showed *CRLF2* deletion (Xp22.33) in addition to two other mutations (*FAT1* deletion (4q35.2), *NUP93* S654G).

#### Management and outcomes

The management of neuroblastoma varied according to extent of disease. Sorrentino and colleagues [6] reported a cohort of 16 patients: 4 patients were stage 1 or 2 and all underwent surgery except for 1 patient who also had chemotherapy; 6 patients were stage 3, and underwent various combinations of surgery, chemotherapy, and radiation, one of whom underwent surgery only but had bone relapse at 5 months; 6 patients were stage 4, all of whom received chemotherapy, 3 of whom also received surgery and 2 also received radiation [6]. Suzuki and colleagues [7] reported a series of 44 patients. Eight patients had locoregional neuroblastoma (five stage 1, one stage 2, five stage 3). According to International Neuroblastoma Response Criteria [A41]: all (5/5) patients with stage 1 disease had complete response to induction therapy; the only patient (1/1) with stage 2 disease had partial response; 80% (4/5) of stage 3 patients had complete response and 20% (1/5) had progressive disease; 21% (7/33) of stage 4 patients had complete response, 21% (7/33) had partial response, 36% (12/33) had no response, and 21% (7/33) had progressive disease [7].

Polishchuk and colleagues [3] reported a cohort of 16 adult patients with recurrent or refractory neuroblastoma managed with ^131^I-MIBG monotherapy. Five of these patients had ganglioneuroblastoma according to Shimada histologic classification [A42]. Treatment toxicities included thrombocytopenia in 69% (20/29; 4 patients grade 3, 16 patients grade 4) [A43]; neutropenia in 69% (20/29; 7 patients grade 3; 13 patients grade 4); need for granulocyte colony-stimulating factor support in 45% (13/29); need for autologous hematopoietic stem cell support in 25% (7/28); 13% (2/15) late-onset grade 2 thyroid abnormalities; 6% (1/16) had second hematologic malignancies. Stiefel and colleagues [10] reported 14 adult neuroblastoma patients with somatic *ALK* mutations, 7 of whom were treated with FDA-approved *ALK* inhibitors.

Adverse events included nausea/vomiting (86%, 6/7) and neurologic side effects (43%, 3/7; hallucinations [1/3], drowsiness [1/3], dizziness [1/3]). Median overall survival since ALK inhibitor treatment was 46.5 (range 17-74) months.

## Discussion

This scoping review provides a detailed overview of adult neuroblastoma cases published in the literature from 1927 to present. We provide several examples of the heterogeneous nature of neuroblastoma, such as an 86-year-old presenting with a mediastinal mass. Our total count of 679 cases provides the literature with an update, as previous literature reviews quoted numbers ranging up to 200 cases. In addition, we detail patient survival according to modern risk stratification guidelines, and introduce recently published outcomes for targeted therapies.

In pediatrics, a considerable proportion of neuroblastoma cases have been associated with mutations of the *ALK* or *PHOX2B* gene [44,45], with somatic acquisitions *ALK* in up to 15% of cases [46]. *MYCN* amplification is reported in approximately 20% of pediatric neuroblastoma patients [44], and has been used as a biomarker to risk stratify patients for poorer prognosis. In our review, we identified seven studies detailing *MYCN* amplification testing, which reported 0 to 14% [7,33] of patients in respective series to test positive for *MYCN* amplification. In fact, the majority of studies reported 0% of patients harboring such a mutation [3,7,29,31,36]. However, Suzuki and colleagues [7] identified that somatic mutations in *ALK* occurred in 42% of patients, and *ATRX* mutations or deletions occurred in 58%. Accordingly, adult neuroblastoma cases may be more associated with progressive accumulation of somatic mutations [7,47], whereas childhood neuroblastoma cases may be more related to genetic susceptibility [44].

Studies of neuroblastoma in pediatrics versus adults show conflicting conclusions [4,7]. Conter and colleagues report in a conference abstract from University of Texas MD Anderson Cancer Center (1994-2012) of 118 adult and 112 pediatric patients no significant difference in survival for L1- (p=0.4), L2- (p=0.54), and M-stage (p=0.73) disease [4]. Esiashvilli and colleagues using SEER data reported a 5-year overall survival to be 84.6% in infants, 47.8% in children 1-9 years old, 46.2% in children 10-19 years old, and 36.3% in adults aged ≥20 years [8]. Navalkee and colleagues also used SEER data (1975-2006) and showed a decreasing survival rate from patients aged 0-4 years to patients aged 15-19 years; there were too few patients in age groups 20-24 and 25-29 years to produce stable survival rates [9]. Accordingly, whether the true survival of adult neuroblastoma patients is inferior to pediatric patients or whether the overall differences are secondary to different disease stage distributions of each population is unknown; however, the consistent reduction in overall survival in the aforementioned studies suggests that adults generally fare worse.

The history of adult neuroblastoma has improved due to advancements of diagnostic and therapeutic techniques. Our literature search spanning nearly a century demonstrates the progression of initially relying solely on basic imaging techniques such as radiography [A4,A6] to routinely using magnetic resonance imaging and MIBG (iodine-123 labeled with metaiodobenzylguanidine) [34,A74]. The use of targeted therapies has also expanded, with studies reporting immunotherapy such as anti-GD2 [7], which was previously used exclusively in high risk children [48]. Future studies would contribute to the literature by providing outcomes of additional adult patients undergoing targeted therapies.

This scoping review has several strengths. First, our literature search was thorough given several methods of identifying articles: searching four databases, reviewing literature searches of previous reviews, and reviewing the references list of all 142 included studies. This increases the confidence of readers that the included number of cases approaches the true number of published cases of adult neuroblastoma. This new count of 679 cases can be quoted in the future to provide clinicians a more accurate impression of the probabilities of encountering this rare entity.

Second, this review outlined the presentations, management strategies, and outcomes using cohort studies. Cohort studies generally provide a more balanced representation of the average adult neuroblastoma case. This contrasts with case reports, which have the aim of showcasing unusual presentations and outcomes. Using our scoping review, readers can appreciate both the general prognosis of adult neuroblastoma and how it varies according to risk stratification, as well as unusual cases we described in our table of case reports.

Nevertheless, this review is not without limitations. As thorough as literature searches may be, they are inherently imperfect, so some cases may have been missed. Second, we were unable to meta-analyze survival data to provide updated estimates. This was due to a lack of individual patient data, as well as suspected difficulty in ascertaining whether patient stages were similarly categorized. Third, there is a potential for overlapping cases if case reports were published both by themselves as well as included in neuroblastoma registries. We aimed to overcome this issue with case series and cohort studies by reviewing the data sources such as the SEER registry. Our total count included only the studies with the highest number of patients of registries to avoid patient duplication. Whereas we agree that some case reports may not be representative of the typical adult neuroblastoma patient, neuroblastoma is a disease regarded for its clinical heterogeneity, so awareness of such obscure cases broadens the differential of the clinician met with this enigmatic disease.

## Conclusion

This scoping review documents the variable presentations, management strategies, and outcomes of 679 adult patients with neuroblastoma. Adult neuroblastoma is a rare disease entity that may present at any age, and appears to show considerable rates of somatic mutations. Future studies evaluating targeted therapies in larger samples are needed.

## Data Availability

All data produced in the present study are available upon reasonable request to the authors

## Acknowledgements

none.

## Author contributions

BHA, BA, AA, AK conceived study idea. AK designed the study protocol. BHA and AK were involved in data extraction and synthesis of results. All authors were involved in manuscript writeup and critical review of the manuscript.

---

**Table A1:** Sample search strategy using medline database.

|  |  |
| --- | --- |
| Database: Embase <1974 to 2022 October 04>, OVID Medline Epub Ahead of Print, In-Process & Other Non-Indexed Citations, Ovid MEDLINE(R) Daily and Ovid MEDLINE(R) 1946 to Present |  |
| Search Strategy: |  |
| 1 | neuroblastoma.ti,ab. (86935) |
| 2 | adult*.ti,ab. (3411525) |
| 3 | geriatrics/ (63893) |
| 4 | geriatric*.ti,ab. (142867) |
| 5 | octogenarian.ti,ab. (3901) |
| 6 | nonagenarian.ti,ab. (1547) |
| 7 | 2 or 3 or 4 or 5 or 6 (3565808) |
| 8 | 1 and 7 (4349) |
| 9 | limit 8 to english language (4111) |
| 10 | limit 9 to humans (3221) |
| 11 | limit 10 to "all adult (19 plus years)" [Limit not valid in Embase; records were retained] (2531) |
| 12 | limit 11 to animal studies [Limit not valid in Ovid MEDLINE(R),Ovid MEDLINE(R) Daily Update,Ovid MEDLINE(R) PubMed not MEDLINE,Ovid MEDLINE(R) In-Process,Ovid MEDLINE(R) Publisher; records were retained] (763) |
| 13 | 11 not 12 (1768) |
| ***** |  |

**Table A2:**
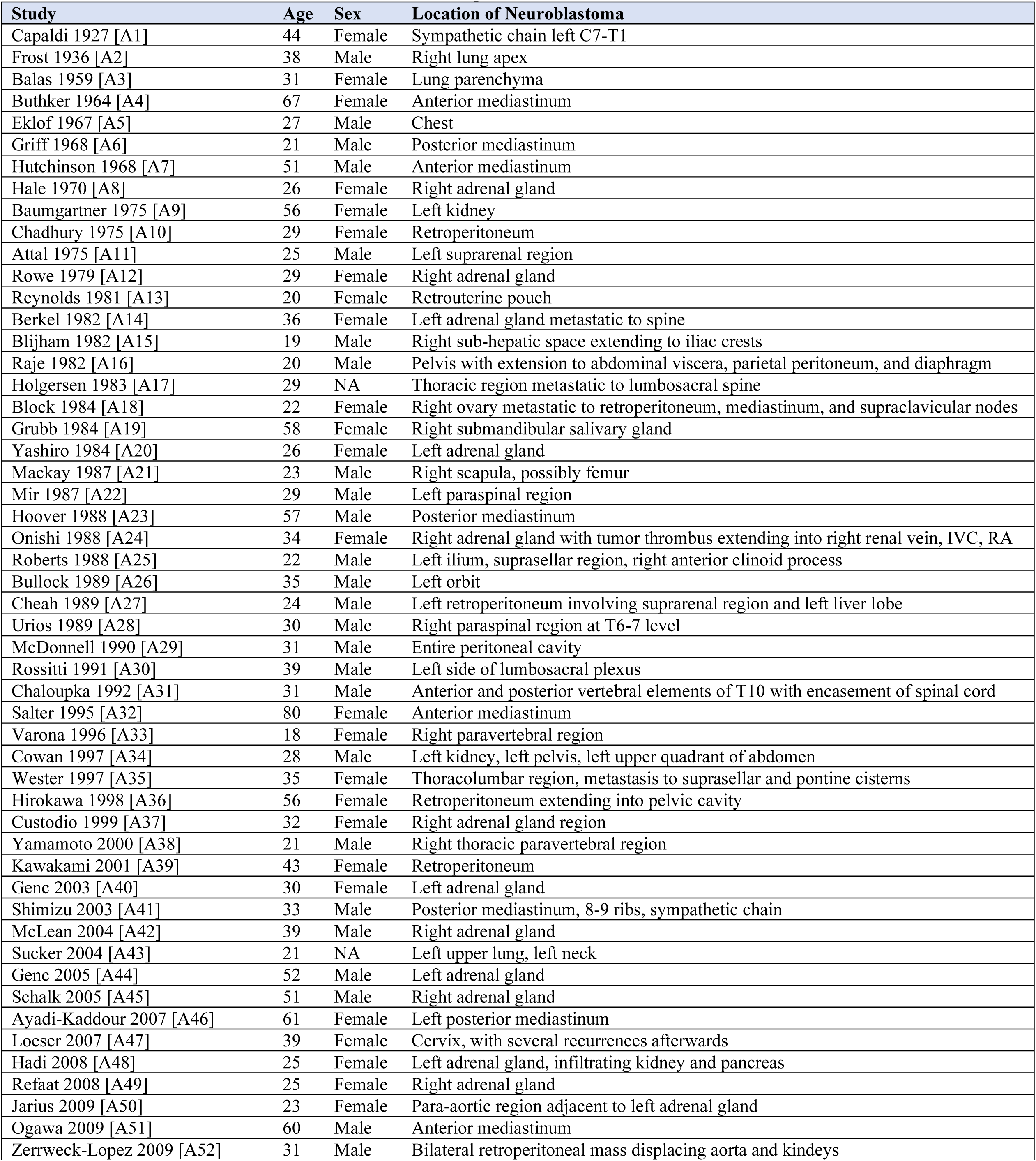
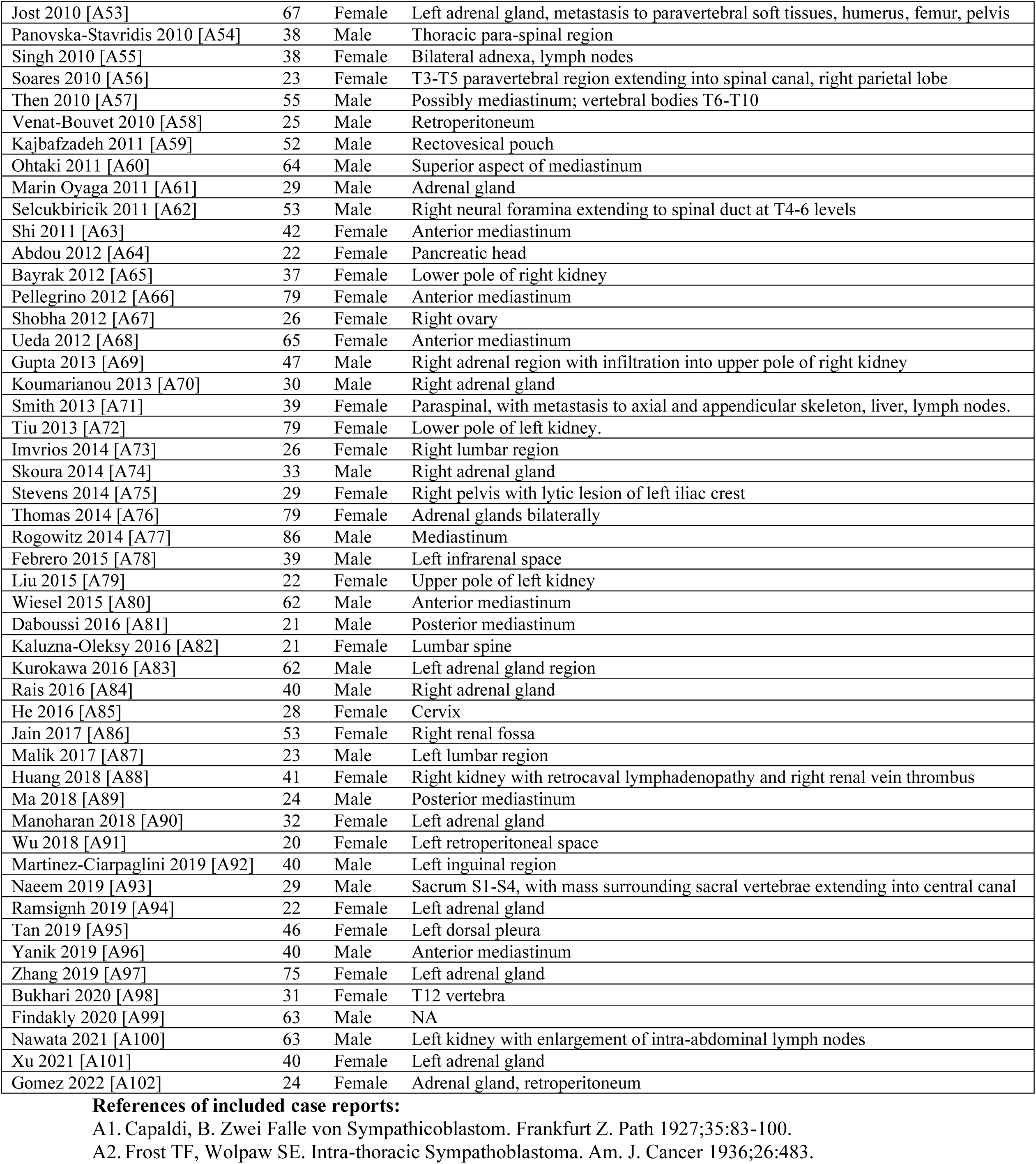
Characteristics of 102 included case reports.

